# Household solid-fuel cooking and visual difficulty in Mexico: a nationally representative cross-sectional study with a negative control outcome

**DOI:** 10.64898/2026.11.26.26369094

**Authors:** Martín Roberto Basile-Alvarez, Enrique C. Guerra, Andrea Malagón-Liceaga, Miguel Ángel Castro-Monreal, Erick André Escalante-Buendía, María Jimena Salas-Muñoz, Mónica Solbes-Gochicoa

## Abstract

**Introduction:** Global burden estimates attribute part of the cataract burden to household air pollution from solid cooking fuels, but most of the evidence comes from South Asia and China, and solid-fuel use is closely tied to poverty. We assessed the association between household solid-fuel cooking and self-reported difficulty seeing among adults in Mexico, using difficulty hearing as a negative control outcome.

**Methods:** We pooled the 2022–2025 rounds of Mexico’s continuous National Health and Nutrition Survey (ENSANUT Continua) for adults aged 40 years or older and linked reported difficulty seeing (with glasses, if worn), glasses use and difficulty hearing to the household’s main cooking fuel, stove type and place of cooking. Survey-weighted quasi-Poisson regression estimated prevalence ratios (PRs) with 95% confidence intervals (CIs). Model 1 was adjusted for age, sex and survey round; the main model (Model 2) also included socioeconomic and health covariates.

**Results:** Of 25,610 adults, 3,465 (10.8% weighted) lived in households that cooked mainly with firewood or charcoal. Any difficulty seeing was reported by 42.9% of adults in solid-fuel households and 33.5% of those in clean-fuel households (Model 1 PR 1.29, 95% CI 1.19 to 1.39). In Model 2, the PR was 1.03 (95% CI 0.92 to 1.15) for any difficulty seeing and 0.94 (95% CI 0.67 to 1.31) for a lot of difficulty or inability to see. For difficulty hearing, the PR was 1.17 (95% CI 1.03 to 1.33) in Model 1 and 1.00 (95% CI 0.84 to 1.19) in Model 2. Stove type and cooking arrangements showed no clear association with difficulty seeing. Glasses use was 27.6% in solid-fuel households and 52.1% in clean-fuel households (Model 2 PR 0.78, 95% CI 0.70 to 0.86).

**Conclusions**

Differences in reported difficulty seeing and hearing between solid-fuel and clean-fuel households narrowed after socioeconomic adjustment. Lower glasses use persisted after adjustment and may reflect unmet need for refractive correction. Assessment of cataract requires measured visual acuity and lens examination.

## Introduction

An estimated 2.1 billion people cook with open fires or inefficient stoves [1], and household air pollution from solid cooking fuels was estimated to cause 3.11 million deaths worldwide in 2021 [2]. The Global Burden of Disease Study counts cataract among the outcomes of household air pollution, with a relative risk of 2.52 (95% confidence interval (CI) 1.36 to 4.50) for solid-fuel use, and attributed 1.96 million years lived with disability from cataract to household air pollution in 2021 [2]. Cataract was the leading cause of blindness in 2020, affecting 15.2 million adults aged 50 years or older, and accounted for 35.4% of age-standardized blindness in Latin America and the Caribbean [3].

The evidence linking solid fuels to eye disease comes largely from South Asia and China. A 2025 meta-analysis of 28 studies reported pooled odds ratios of 2.29 (95% CI 1.24 to 4.23) for cataract and 1.70 (95% CI 1.45 to 2.00) for vision impairment, with very high heterogeneity for cataract [4]. Among 486,532 Chinese adults, long-term solid-fuel use was associated with incident cataract identified through health-insurance records (odds ratio 1.17, 95% CI 1.08 to 1.26) [5], and among Chinese adults aged 45 years or older it was associated with incident self-reported distance vision impairment (hazard ratio 1.38, 95% CI 1.28 to 1.50) [6]. Because solid fuels are used mostly by poorer and rural households, confounding is a central concern. In a national survey in India with measured visual acuity, the excess prevalence of vision impairment among older adults using unclean fuels fell from 9.0 to 3.2 percentage points after socioeconomic adjustment [7]. Women, who spend the most time near the domestic hearth, are often more exposed [1], and some studies have reported associations with cataract mainly in women [5,8], although the sex-specific estimates pooled in the 2025 meta-analysis were similar in men and women [4]. The only estimate for Mexico, from the Mexican sample of the World Health Organization Study on global AGEing and adult health, was an odds ratio for cataract of 1.23 (95% CI 0.84 to 1.81) [4]. Apart from that multicountry survey, we found no study of solid-fuel cooking and vision in Mexico or elsewhere in Latin America, and none that used a negative control outcome.

In Mexico, 9.7% of dwellings used firewood or charcoal for cooking in 2025, rising to 35.9% in localities with fewer than 2,500 inhabitants [9]. Firewood is often used together with liquefied petroleum (LP) gas, so the main fuel reported in surveys captures only part of the exposure [10]. In rural homes in the state of Michoacán, open wood fires produced a 48-hour mean fine particulate matter concentration of 693 μg/m³ near the stove [11], and in a randomized trial in the same state, women who used an improved biomass stove most of the time reported less eye discomfort than those who cooked over an open fire [12]. Wood use as the main cooking fuel is concentrated in rural households of low socioeconomic position [13], and low socioeconomic position is itself associated with vision impairment in Mexico [14].

Mexico’s continuous National Health and Nutrition Survey (Encuesta Nacional de Salud y Nutrición Continua, ENSANUT Continua) records the main cooking fuel, stove and place of cooking of each household, and asks adults about difficulty seeing and hearing with the questions of the Washington Group on Disability Statistics, which have been used to describe severe vision impairment in Mexico [14]. A module with measured visual acuity and assessment of media opacity was added in 2025 [15], but its data were not publicly available at the time of this analysis. Because solid-fuel use is so closely tied to poverty, we used difficulty hearing as a negative control outcome. Difficulty hearing shares potential confounders with difficulty seeing, such as age, poverty, rurality and access to care, but is not thought to be affected by smoke through the same pathway, so associations of similar magnitude with both outcomes would be consistent with residual confounding or reporting bias [16]. We estimated the association of household solid-fuel cooking with difficulty seeing among adults aged 40 years or older, examined stove type, place of cooking, sleeping where the household cooks and modification by sex, and described glasses use.

## Materials and Methods

### Study design and data sources

ENSANUT Continua is a series of annual cross-sectional household surveys in Mexico. Each round uses an independent probability sample with a stratified cluster design and its own expansion weights, strata and primary sampling units (PSUs) [17,18]. We used the public-use files of the household questionnaire, the household roster and the adult questionnaire (adults aged 20 years or older) of each round, with their catalogs and questionnaires, released by the Instituto Nacional de Salud Pública (INSP) [19]. An informant answers the household questionnaire and roster for the whole household, and each selected adult answers the adult questionnaire. The main analysis pooled the 2022–2025 rounds, for which INSP released a household wellbeing index; the 2021 round, which lacks that index, was used in a sensitivity analysis. The 2024 and 2025 files contain no records from the state of Sinaloa. Reporting follows the Strengthening the Reporting of Observational Studies in Epidemiology (STROBE) recommendations for cross-sectional studies, and the completed checklist is provided in the Appendices [20].

### Participants

We included adults aged 40 years or older, because cataract and presbyopia are age-related [21], who answered the seeing items and lived in households that cooked at home with solid or clean fuel. All adults were asked the seeing items in 2022 and 2025. In 2023 and 2024, the adult questionnaire assigned one half of the selected adults aged 60 years or older to the functioning section, which includes the questions on difficulty seeing, and the other half to a section on basic activities of daily living. The weights of those who answered the functioning section were multiplied by the ratio of the weighted total of all adults of that age to the weighted total of those who answered it (2.05 in 2023 and 2.06 in 2024).

### Cooking fuel and cooking arrangements

The household questionnaire asks which fuel the household uses most for cooking: firewood, charcoal, LP gas, natural gas, electricity, another fuel, or none. We compared households that used solid fuel (firewood or charcoal) with those that used clean fuel (LP gas, natural gas or electricity) and excluded households that did not cook at home or used another fuel. Among solid-fuel households, we grouped the reported stove type as an open fire or oven without a chimney or hood; an open fire or oven with a chimney or hood, or a closed oven with a chimney; or a gas or electric stove (suggesting use of more than one fuel) or another, unspecified stove. The questionnaire asks whether the dwelling has a room for cooking. If it does, it asks whether household members also sleep there; if not, it asks whether the household cooks in a corridor, under a small roof or outdoors. Wording and codes were the same in all rounds (Appendices).

### Outcomes

The functioning section uses questions of the Washington Group on Disability Statistics [22]. Adults were asked whether they wear glasses or contact lenses; wearers were asked whether they have difficulty seeing when wearing them, and the others whether they have difficulty seeing, with four response options: no difficulty, some difficulty, a lot of difficulty, or cannot see at all. The primary outcomes were any difficulty seeing (some difficulty or more) and a lot of difficulty or inability to see. Glasses use was a secondary outcome that described the use of refractive correction; the survey did not measure the need for correction. Difficulty hearing (some difficulty or more, with a hearing aid if used), asked in the same way, was the negative control outcome. Both are self-reported functional difficulties that share determinants such as age, poverty, rurality, access to care and a tendency to report difficulties, which negative controls can also detect [16], whereas cataract, but not hearing loss, is among the outcomes attributed to household air pollution in burden estimates [2]. Because solid-fuel use has been associated with hearing loss in Chinese cohorts [23], the negative control is imperfect: similar associations with both outcomes would be compatible with shared bias or with effects on both senses.

### Covariates

Model 1 was adjusted for age (a natural cubic spline with knots at 49 and 61 years, the weighted tertiles of the analyzed adults), sex and survey round. Model 2, the main model, added schooling, household wellbeing quintile, rural residence (localities with fewer than 2,500 inhabitants), region (the eight regions of the survey [17]) and speaking an Indigenous language, which may influence both the fuel a household uses and difficulty seeing or its report [13,14], as well as diagnosed diabetes and smoking (current, former or never), which are risk factors for cataract [24,25] and differ by socioeconomic position. Because household air pollution has also been linked to type 2 diabetes [2], diabetes could lie partly on the causal path, so we also report the model before diabetes and smoking were added.

The INSP household wellbeing index is the first principal component of the polychoric correlations of eight household characteristics, one of which counts goods that include a stove [26]. In the household files, this item is ownership of a gas stove, reported by 97% to 98% of clean-fuel households but by only 48% to 60% of solid-fuel households. Adjusting for it would partly adjust for a close proxy of the exposure and remove part of the contrast of interest. We therefore rebuilt the index in each round from all households in the household file, once with the eight variables, to check the reconstruction, and once without the gas stove. We used quintiles of the second version, cut at unweighted quintiles of the households of each round, the rule that reproduces the released quintiles (Appendices).

## Statistical analysis

All analyses accounted for the survey design, with PSUs nested within strata, using the survey package in R. Pooled weights divided each annual weight by the number of pooled rounds in which the adult’s state had records. The design was formed from all adult records, and the analyzed adults were treated as a domain of it. Strata with a single PSU (53 of 337), and strata left with a single PSU in a domain, were handled by centering at the grand mean.

Survey-weighted quasi-Poisson models with a log link estimated prevalence ratios (PRs), with confidence intervals based on the t distribution with the residual degrees of freedom of each model [27]. We used predictive margins to estimate prevalence standardized to the covariate distribution of the analyzed adults. The two primary outcomes formed one family for Holm correction, as did the contrasts with clean fuel within each analysis of cooking arrangements; other p values are unadjusted. To compare difficulty seeing formally with the negative control, we fitted one model with one record per adult per outcome and interacted every term with outcome type. Exponentiating the fuel-by-outcome interaction gives the ratio of the PR for difficulty seeing to that for difficulty hearing, and the linearized variance accounts for the paired records. To show which covariates accounted for the difference between Models 1 and 2, we added them to Model 1 in groups.

Using Model 2, we compared stove type, place of cooking and sleeping where the household cooks with clean fuel, using a Wald test across groups and a contrast within solid-fuel households. For stove type, we also tested for trend by scoring clean fuel and the three solid-fuel stove groups. We tested effect modification by sex as planned, given women’s greater role in cooking, and explored modification by age group (40–59 years and 60 years or older) and by residence. Each interaction was added separately to Model 2, and subgroup PRs were estimated from those models. For comparison, we also fitted Model 2 separately in each subgroup.

Sensitivity analyses used average-stratum variances; restricted the data to the 31 states with records in every round; replaced the rebuilt wellbeing quintile with the quintile released by INSP or with the index rebuilt with all eight variables, or removed it (in both the 2022–2025 and the 2021–2025 data); added glasses use or diagnosed hypertension; excluded adults with diagnosed diabetes; excluded clean-fuel households that heat the dwelling with solid fuel, solid-fuel households that reported a gas, electric or other stove, or charcoal users; and capped the weights at the 99th percentile of each round. Consistency across rounds was tested with a product term of fuel by round. E-values were calculated for Model 2 [28]. Only smoking status had missing values, for 85 adults (0.3% weighted), so analyses were complete-case. Coefficients of covariates are reported for transparency only. As a check of the data preparation, we reproduced the published 2023 estimates of households cooking with firewood or charcoal and of households without a room for cooking [29].

## Ethics, data access and reproducibility

The analyses used public-use, de-identified files, and the authors had no access to identifying information. No additional institutional review was sought for this secondary analysis of publicly available data. ENSANUT publications describe the ethical oversight of the source surveys: the 2022 methodology report states that the INSP Ethics, Research and Biosafety Committees approved the questionnaires, interview procedures and consent forms and that participants gave informed consent [17], and the 2025 report states that the protocol was approved by the INSP Ethics and Biosafety Committees and that all participants gave informed consent [18]. The workflow was run in R version 4.3.3 (R Foundation for Statistical Computing, Vienna, Austria), with tidyverse packages and psych (polychoric correlations) for data preparation, survey (version 4.5) and splines for survey-weighted analyses, ggplot2 for figures and flextable for tables. The analysis scripts record the versions of all required packages.

## Results

### Participants and cooking conditions

The 2022–2025 adult files contained 44,223 records, including 28,361 adults aged 40 years or older. Of these, 2,422 adults aged 60 years or older had been assigned to the section on basic activities of daily living and were not asked about difficulty seeing. We excluded a further 329 adults because their household did not cook at home, used another fuel or lacked data on cooking, or because their smoking status was missing. The analysis included 25,610 adults, of whom 3,465 (10.8% weighted) lived in households that cooked mainly with solid fuel (Figure 1; Appendices, Table 7). The design included 337 strata and 1,549 PSUs.

**Figure 1:**
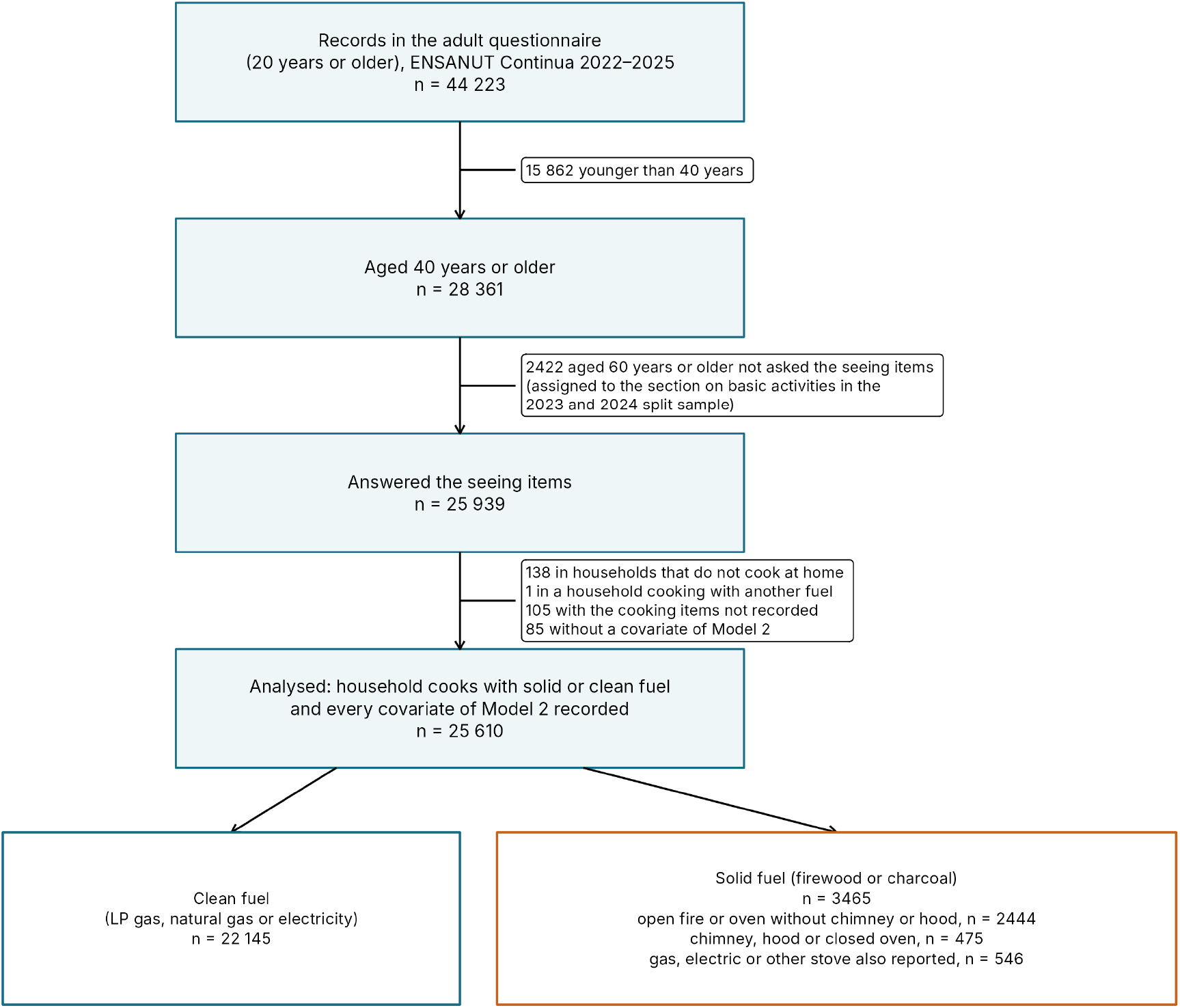
Selection of adults aged 40 years or older, ENSANUT Continua 2022–2025. In 2023 and 2024, the adult questionnaire assigned one half of the selected adults aged 60 years or older to the functioning section, which holds the questions on glasses and difficulty seeing, and the other half to the section on basic activities of daily living. Cooking fuel and stove type come from the household questionnaire (items h0109 and h0110). Model 2 covariates: age, sex, survey round, schooling, household wellbeing quintile, rural residence, region, Indigenous language, diagnosed diabetes and smoking. ENSANUT: Encuesta Nacional de Salud y Nutrición (National Health and Nutrition Survey); LP: liquefied petroleum.

In the household files, the weighted proportion of households cooking mainly with firewood or charcoal ranged from 9.6% to 11.6% across rounds and was 34.9% in rural and 3.9% in urban households (Appendices, Table 8). For 2023, our estimates reproduced the published proportions of households cooking with firewood or charcoal (9.6%) and of households without a room for cooking (3.5%, 95% CI 2.4 to 4.9) [29]. Among solid-fuel households, 65% to 74% burned the fuel in an open fire or oven without a chimney or hood. The index rebuilt without the gas stove correlated with the released index at 0.967 to 0.979, and unweighted cut points applied to the released index reproduced 97.5% to 99.6% of the released quintiles (Appendices, Table 9).

Adults in solid-fuel households were of similar age to those in clean-fuel households but more often lived in rural areas (67.9% vs 14.1%), spoke an Indigenous language (32.0% vs 2.6%), had no schooling (20.4% vs 5.2%) and belonged to the lowest wellbeing quintile (67.7% vs 11.6%); they less often reported diagnosed diabetes and current smoking (Table 1). Most lived in dwellings with a room for cooking (85.1%), and 4.5% lived in households whose members slept where they cooked.

**Table 1:** Characteristics of adults aged 40 years or older by the main cooking fuel of the household, ENSANUT Continua 2022–2025.

| Characteristic | Category | All adults | Clean fuel | Solid fuel |
|---|---|---|---|---|
| Adults, n (weighted %) | All | 25,610 (100.0) | 22,145 (89.2) | 3,465 (10.8) |
| Age in years, mean (SD) | All | 56.3 (12.0) | 56.3 (11.9) | 56.4 (12.6) |
| Age group (years) | 40–59 | 16,177 (63.3) | 14,007 (63.2) | 2,170 (64.2) |
|  | 60 or older | 9,433 (36.7) | 8,138 (36.8) | 1,295 (35.8) |
| Women | All | 15,000 (52.7) | 13,050 (53.1) | 1,950 (49.1) |
| Rural residence | All | 6,620 (19.9) | 4,222 (14.1) | 2,398 (67.9) |
| Region | Pacific North | 3,684 (9.4) | 3,494 (10.2) | 190 (2.6) |
|  | Border | 3,730 (12.0) | 3,661 (13.3) | 69 (1.7) |
|  | Pacific-Center | 1,877 (11.1) | 1,744 (11.9) | 133 (4.8) |
|  | North-Center | 5,229 (12.9) | 4,817 (13.6) | 412 (7.8) |
|  | Center | 2,369 (10.1) | 1,944 (9.2) | 425 (16.9) |
|  | Mexico City/State of Mexico | 2,003 (22.7) | 1,950 (24.8) | 53 (5.2) |
|  | Pacific South | 3,140 (12.4) | 2,118 (9.7) | 1,022 (35.0) |
|  | Peninsula | 3,578 (9.4) | 2,417 (7.4) | 1,161 (26.0) |
| Speaks an Indigenous language | All | 1,894 (5.8) | 677 (2.6) | 1,217 (32.0) |
| Schooling | No schooling | 2,074 (6.9) | 1,336 (5.2) | 738 (20.4) |
|  | Primary | 8,781 (30.9) | 6,967 (28.4) | 1,814 (51.7) |
|  | Secondary | 7,359 (28.6) | 6,625 (29.4) | 734 (22.3) |
|  | Upper secondary or higher | 7,396 (33.6) | 7,217 (37.0) | 179 (5.5) |
| Household wellbeing quintile | Q5 (highest) | 5,371 (24.9) | 5,321 (27.7) | 50 (1.3) |
|  | Q4 | 4,957 (19.7) | 4,854 (21.7) | 103 (2.7) |
|  | Q3 | 5,019 (19.9) | 4,780 (21.5) | 239 (6.4) |
|  | Q2 | 5,117 (17.9) | 4,327 (17.4) | 790 (21.8) |
|  | Q1 (lowest) | 5,146 (17.7) | 2,863 (11.6) | 2,283 (67.7) |
| Diagnosed diabetes | All | 5,100 (18.6) | 4,490 (18.9) | 610 (16.3) |
| Diagnosed hypertension | All | 7,393 (27.2) | 6,604 (27.7) | 789 (22.8) |
| Smoking | Never | 17,533 (65.1) | 14,904 (64.2) | 2,629 (72.5) |
|  | Former | 4,560 (19.1) | 4,057 (19.4) | 503 (16.0) |
|  | Current | 3,517 (15.9) | 3,184 (16.4) | 333 (11.5) |
| Where the household cooks | Room for cooking | 24,646 (96.7) | 21,674 (98.1) | 2,972 (85.1) |
|  | Corridor or small roof | 844 (2.9) | 445 (1.8) | 399 (12.3) |
|  | Outdoors | 120 (0.4) | 26 (0.1) | 94 (2.6) |
| Household members sleep where they cook | All | 1,207 (4.2) | 986 (4.2) | 221 (4.5) |
| Dwelling heated with solid fuel | All | 672 (2.1) | 522 (1.7) | 150 (5.3) |
| Wears glasses or contact lenses | All | 12,534 (49.4) | 11,549 (52.1) | 985 (27.6) |
| Any difficulty seeing | All | 9,515 (34.5) | 8,006 (33.5) | 1,509 (42.9) |
| A lot of difficulty seeing or cannot see | All | 1,223 (4.0) | 974 (3.6) | 249 (6.6) |
| Any difficulty hearing | All | 3,867 (15.4) | 3,275 (15.0) | 592 (18.1) |
| Survey round | 2022 | 7,305 (25.8) | 6,288 (25.7) | 1,017 (27.0) |
|  | 2023 | 3,352 (25.2) | 3,016 (25.4) | 336 (23.8) |
|  | 2024 | 6,320 (24.5) | 5,469 (24.5) | 851 (24.5) |
|  | 2025 | 8,633 (24.5) | 7,372 (24.4) | 1,261 (24.7) |

Values are unweighted n (survey-weighted column %) unless stated otherwise; the first row gives n (weighted % of all analyzed adults). Means and standard deviations (SDs) are survey-weighted; the SD is descriptive. Clean fuel: liquefied petroleum gas, natural gas or electricity; solid fuel: firewood or charcoal (item h0109, the fuel most used for cooking). Difficulty seeing and hearing: some difficulty, a lot of difficulty or cannot, with glasses or a hearing aid if used. Household wellbeing: quintile (Q1, lowest, to Q5, highest) of the Instituto Nacional de Salud Pública index of wellbeing conditions, rebuilt without the gas stove. Pooled weights divide each annual weight by the number of pooled rounds in which the state had records; the weights of adults aged 60 years or older who answered the functioning section in 2023 and 2024 were rescaled. ENSANUT: Encuesta Nacional de Salud y Nutrición (National Health and Nutrition Survey).

### Solid-fuel cooking, difficulty seeing, and the negative control

Any difficulty seeing was reported by 42.9% (95% CI 39.5 to 46.4) of adults in solid-fuel households and 33.5% (95% CI 32.2 to 34.8) of those in clean-fuel households. A lot of difficulty or inability to see was reported by 6.6% and 3.6%, respectively (Table 2; round-specific estimates in Appendices, Table 11). After adjustment for age, sex and survey round (Model 1), the PRs were 1.29 (95% CI 1.19 to 1.39) and 1.76 (95% CI 1.31 to 2.36), respectively. In Model 2, the PRs were 1.03 (95% CI 0.92 to 1.15; p=0.589) and 0.94 (95% CI 0.67 to 1.31; p=0.701). The standardized prevalence of any difficulty seeing was 35.4% (95% CI 31.9 to 39.1) in solid-fuel households and 34.4% (95% CI 33.2 to 35.7) in clean-fuel households, a difference of 1.0 percentage point (95% CI −2.8 to 4.9) (Appendices, Table 12).

**Table 2:**
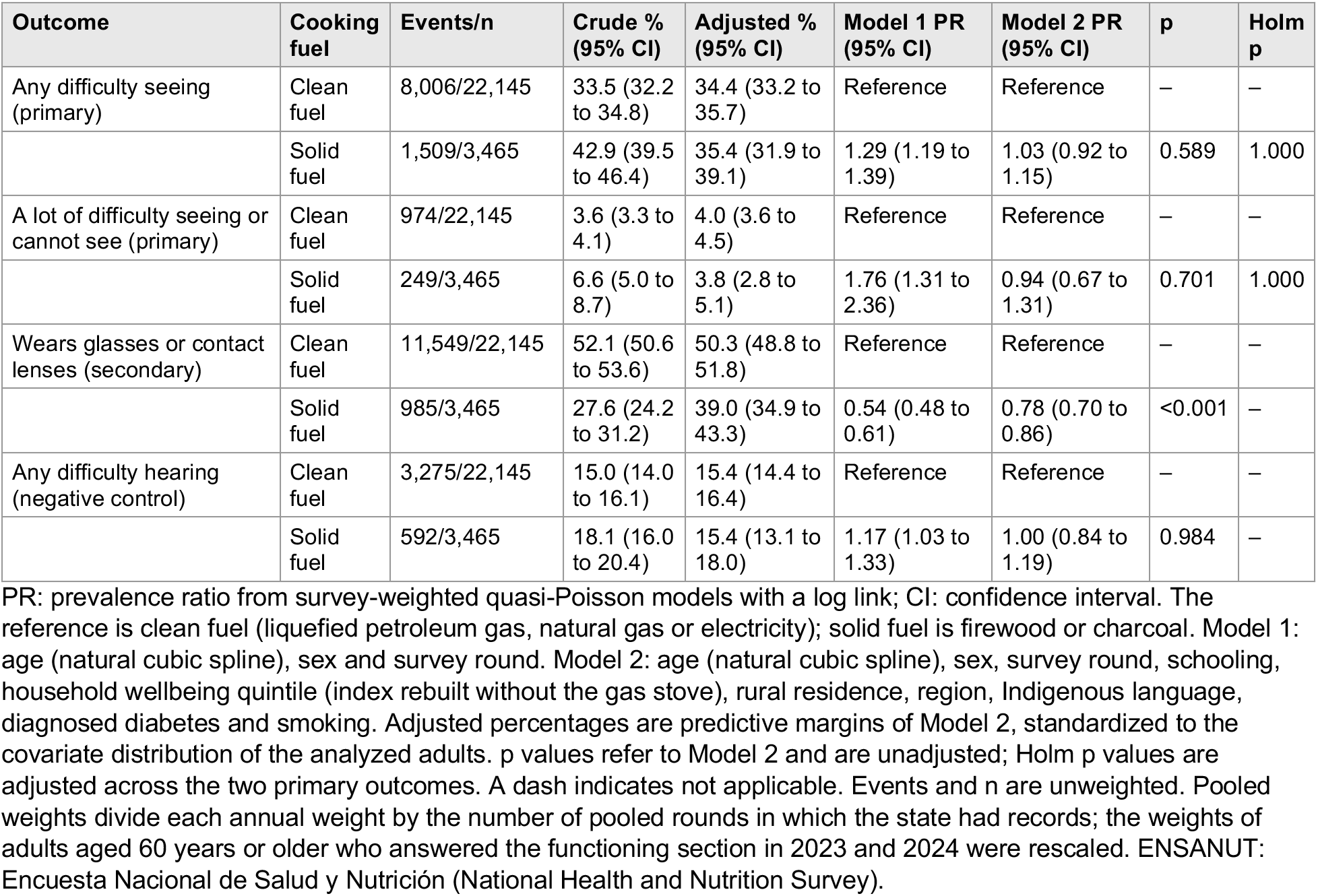
Difficulty seeing, glasses use and difficulty hearing by the main cooking fuel of the household, adults aged 40 years or older, ENSANUT Continua 2022–2025.

Difficulty hearing was reported by 18.1% of adults in solid-fuel households and 15.0% of those in clean-fuel households. The PR was 1.17 (95% CI 1.03 to 1.33) in Model 1 and 1.00 (95% CI 0.84 to 1.19) in Model 2 (Table 2). In the joint model, the ratio of the PR for difficulty seeing to the PR for difficulty hearing was 1.10 (95% CI 0.93 to 1.29; p=0.269) with Model 1 covariates and 1.03 (95% CI 0.83 to 1.27; p=0.795) with Model 2 covariates (Table 3). Adding schooling and household wellbeing to Model 1 brought the PRs to 1.04 (95% CI 0.94 to 1.15) for difficulty seeing and 0.98 (95% CI 0.86 to 1.13) for difficulty hearing, and further adjustment changed them little. Before diabetes and smoking, which could lie partly on the causal path, were added, the PR for difficulty seeing was 1.03 (95% CI 0.92 to 1.15) (Figure 2; Appendices, Table 13).

**Figure 2:**
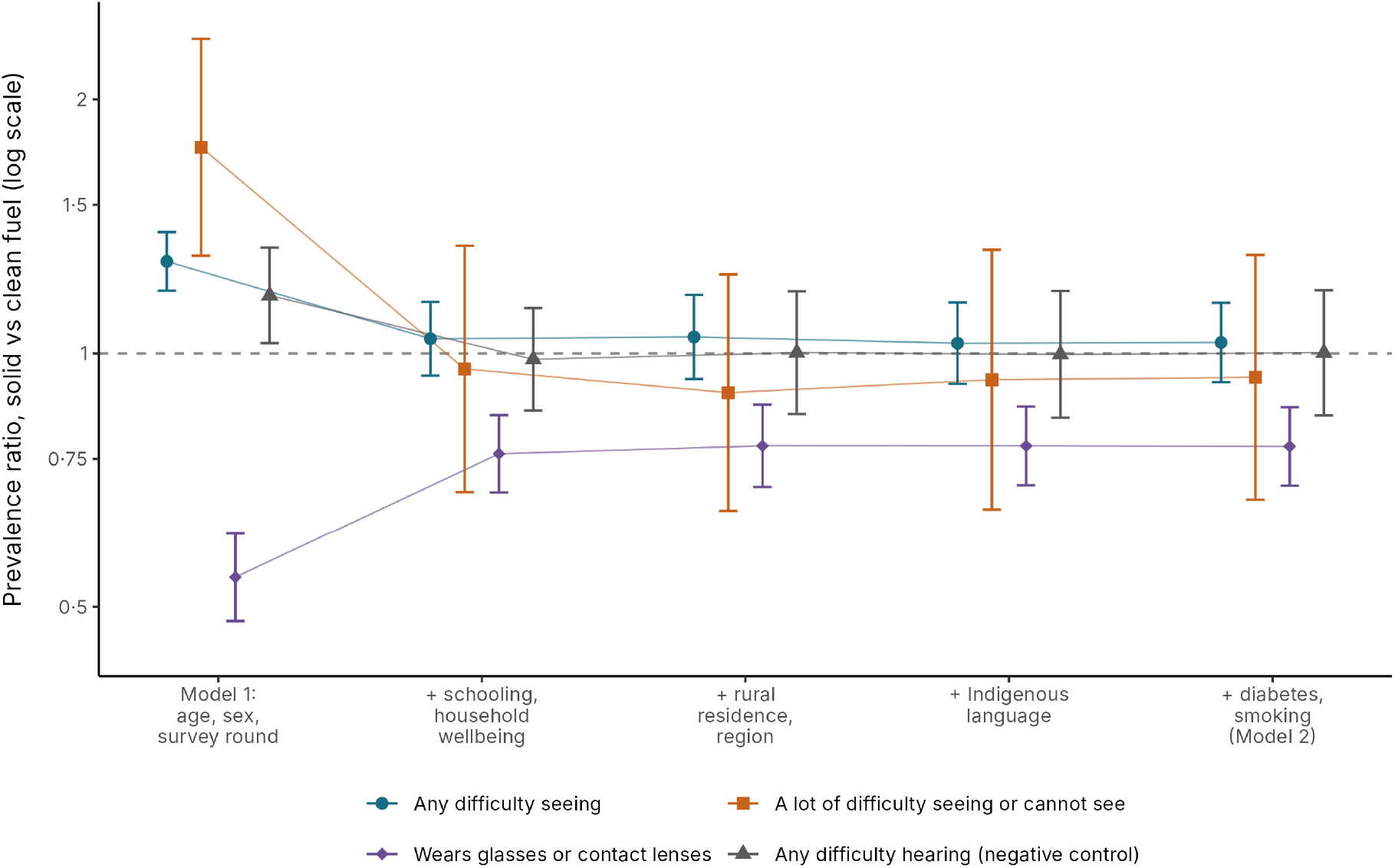
Prevalence ratios for solid vs clean cooking fuel with covariates added in groups. Points show prevalence ratios (logarithmic scale), and bars show 95% confidence intervals, from survey-weighted quasi-Poisson models fitted in the same analyzed adults. Each step adds the covariates named to those of the step on its left, and the last step is Model 2. Difficulty hearing is the negative control outcome.

**Table 3:** Difficulty seeing relative to the negative control outcome, difficulty hearing, adults aged 40 years or older, ENSANUT Continua 2022–2025.

| **Comparison** | **Model** | **Ratio of PRs (95% CI)** | **p** |
|---|---|---|---|
| PR for any difficulty seeing divided by PR for any difficulty hearing, solid vs clean fuel | Model 1 | 1.10 (0.93 to 1.29) | 0.269 |
| PR for any difficulty seeing divided by PR for any difficulty hearing, solid vs clean fuel | Model 2 | 1.03 (0.83 to 1.27) | 0.795 |

Product term of fuel by outcome type in one survey-weighted quasi-Poisson model for the two outcomes, with every covariate interacted with outcome type. Each adult contributes one record per outcome, and the linearized variance accounts for the pairing. Model 1: age (natural cubic spline), sex and survey round. Model 2: Model 1 plus schooling, household wellbeing quintile (index rebuilt without the gas stove), rural residence, region, Indigenous language, diagnosed diabetes and smoking. PR: prevalence ratio; CI: confidence interval; ENSANUT: Encuesta Nacional de Salud y Nutrición (National Health and Nutrition Survey).

Glasses use was reported by 27.6% (95% CI 24.2 to 31.2) of adults in solid-fuel households and 52.1% (95% CI 50.6 to 53.6) of those in clean-fuel households (Table 2). The PR was 0.54 (95% CI 0.48 to 0.61) in Model 1 and 0.78 (95% CI 0.70 to 0.86) in Model 2. The standardized prevalence was 39.0% (95% CI 34.9 to 43.3) in solid-fuel households and 50.3% (95% CI 48.8 to 51.8) in clean-fuel households, a difference of −11.3 percentage points (95% CI −15.5 to −7.0).

### Cooking arrangements

Compared with clean fuel, the PRs for any difficulty seeing ranged from 0.92 to 1.19 across stove types, places of cooking and sleeping arrangements, all with confidence intervals including 1 (Table 4; Figure 3; Appendices, Table 14). Within solid-fuel households, an open fire without a chimney or hood was not associated with more difficulty seeing than a chimney, hood or closed oven (ratio 0.91, 95% CI 0.76 to 1.10), and neither was cooking in a corridor, under a small roof or outdoors rather than in a room (ratio 0.88, 95% CI 0.71 to 1.09) or sleeping where the household cooks (ratio 0.90, 95% CI 0.73 to 1.12). The tests for trend across stove groups gave p=0.749 for any difficulty seeing and p=0.947 for a lot of difficulty or inability to see. For the negative control, difficulty hearing was more common with a chimney, hood or closed oven (PR 1.33, 95% CI 1.01 to 1.75; Holm-adjusted p=0.122) and, imprecisely, in households that sleep where they cook (PR 1.35, 95% CI 0.96 to 1.89), without a consistent pattern across groups (p=0.080 for stove type).

**Figure 3:**
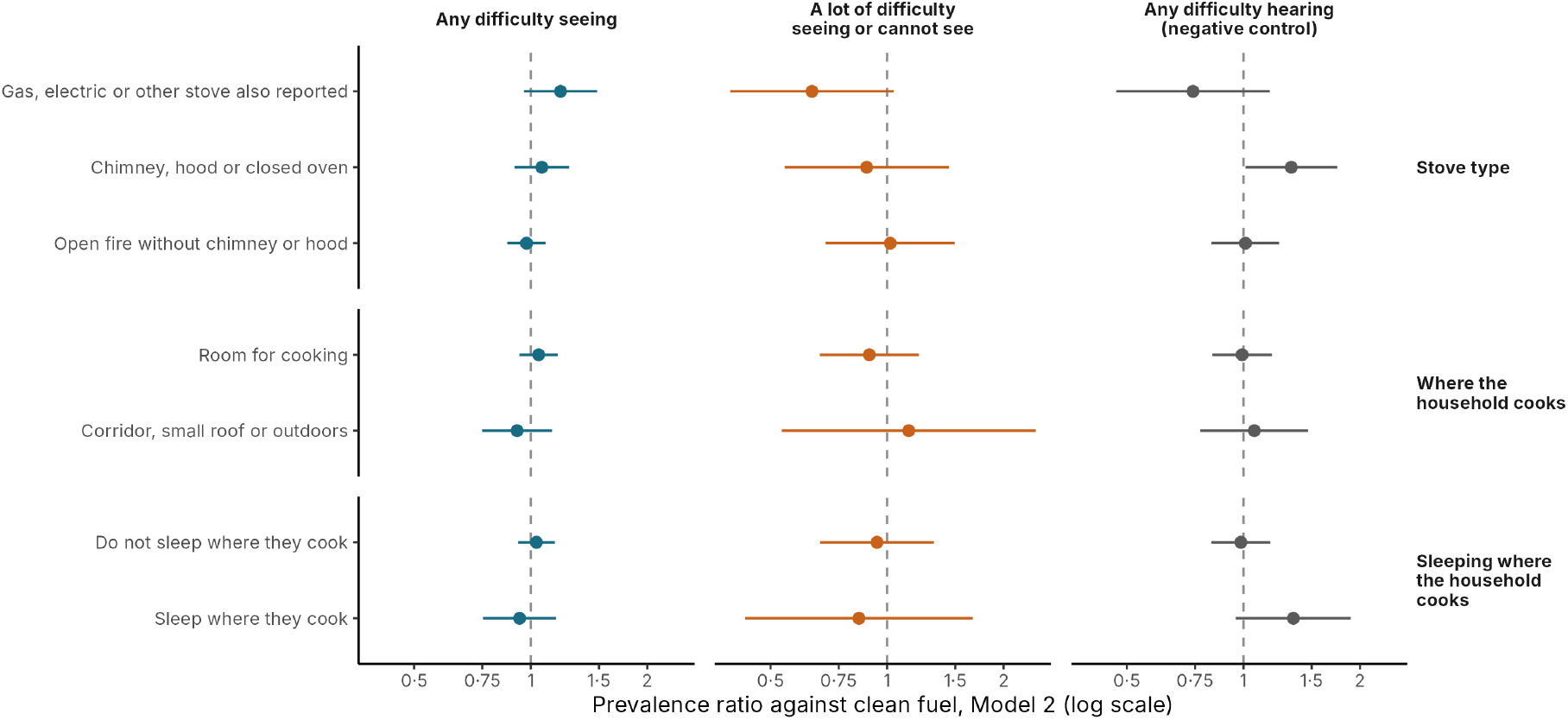
Difficulty seeing and difficulty hearing by stove type, place of cooking and sleeping where the household cooks. Prevalence ratios from Model 2 (logarithmic scale) against households cooking with clean fuel, with 95% confidence intervals. All groups shown cook mainly with firewood or charcoal. Model 2 covariates: age (natural cubic spline), sex, survey round, schooling, household wellbeing quintile (index rebuilt without the gas stove), rural residence, region, Indigenous language, diagnosed diabetes and smoking.

**Table 4:** Difficulty seeing and difficulty hearing by stove type, place of cooking and sleeping where the household cooks, adults aged 40 years or older, ENSANUT Continua 2022–2025.

| Exposure | Group | n | Any difficulty seeing, adjusted % (95% CI) | Any difficulty seeing, PR (95% CI) | A lot of difficulty seeing or cannot see, adjusted % (95% CI) | A lot of difficulty seeing or cannot see, PR (95% CI) | Any difficulty hearing, adjusted % (95% CI) | Any difficulty hearing, PR (95% CI) |
|---|---|---|---|---|---|---|---|---|
| Reference | Clean fuel | 22,145 | 34.4 (33.2 to 35.7) | Reference | 4.0 (3.6 to 4.5) | Reference | 15.4 (14.4 to 16.4) | Reference |
| Stove type | Solid fuel, gas, electric or other stove also reported | 546 | 41.1 (32.7 to 50.0) | 1.19 (0.96 to 1.48) | 2.6 (1.6 to 4.1) | 0.64 (0.39 to 1.04) | 11.4 (7.1 to 17.7) | 0.74 (0.47 to 1.17) |
| Stove type | Solid fuel, chimney, hood or closed oven | 475 | 36.8 (31.1 to 42.8) | 1.07 (0.91 to 1.26) | 3.6 (2.2 to 5.7) | 0.89 (0.54 to 1.44) | 20.4 (15.4 to 26.5) | 1.33 (1.01 to 1.75) |
| Stove type | Solid fuel, open fire without chimney or hood | 2,444 | 33.6 (30.0 to 37.4) | 0.98 (0.87 to 1.09) | 4.1 (2.9 to 5.8) | 1.02 (0.69 to 1.50) | 15.5 (12.9 to 18.7) | 1.01 (0.83 to 1.24) |
| Stove type | Within solid fuel: open fire without chimney or hood vs chimney, hood or closed oven, ratio (95% CI); p | – | – | 0.91 (0.76 to 1.10);<br>p=0.342 | – | 1.15 (0.69 to 1.93);<br>p=0.595 | – | 0.76 (0.54 to 1.07);<br>p=0.116 |
| Stove type | Global test of the groups, p | – | – | 0.203 | – | 0.303 | – | 0.080 |
| Stove type | Trend across the four groups, p | – | – | 0.749 | – | 0.947 | – | 0.666 |
| Place of cooking | Solid fuel, room for cooking | 2,972 | 36.1 (32.2 to 40.1) | 1.05 (0.94 to 1.17) | 3.6 (2.8 to 4.7) | 0.90 (0.67 to 1.21) | 15.2 (12.9 to 18.0) | 0.99 (0.83 to 1.19) |
| Place of cooking | Solid fuel, corridor, small roof or outdoors | 493 | 31.7 (25.7 to 38.4) | 0.92 (0.75 to 1.14) | 4.6 (2.2 to 9.4) | 1.14 (0.53 to 2.42) | 16.4 (11.9 to 22.2) | 1.07 (0.77 to 1.47) |
| Place of cooking | Within solid fuel: corridor, small roof or outdoors vs room for cooking, ratio (95% CI); p | – | – | 0.88 (0.71 to 1.09);<br>p=0.244 | – | 1.26 (0.65 to 2.45);<br>p=0.488 | – | 1.08 (0.78 to 1.49);<br>p=0.661 |
| Place of cooking | Global test of the groups, p | – | – | 0.456 | – | 0.579 | – | 0.907 |
| Sleeping where the household cooks | Solid fuel, do not sleep where they cook | 3,244 | 35.6 (32.0 to 39.4) | 1.03 (0.93 to 1.15) | 3.8 (2.8 to 5.1) | 0.94 (0.67 to 1.32) | 15.1 (12.8 to 17.8) | 0.99 (0.83 to 1.17) |
| Sleeping where the household cooks | Solid fuel, sleep where they cook | 221 | 32.2 (25.7 to 39.4) | 0.94 (0.75 to 1.16) | 3.4 (1.7 to 6.5) | 0.85 (0.43 to 1.67) | 20.7 (14.6 to 28.5) | 1.35 (0.96 to 1.89) |
| Sleeping where the household cooks | Within solid fuel: sleep where they cook vs do not, ratio (95% CI); p | – | – | 0.90 (0.73 to 1.12);<br>p=0.356 | – | 0.90 (0.46 to 1.76);<br>p=0.755 | – | 1.37 (0.97 to 1.92);<br>p=0.073 |
| Sleeping where the household cooks | Global test of the groups, p | – | – | 0.599 | – | 0.866 | – | 0.192 |

Survey-weighted quasi-Poisson models with the Model 2 covariates: age (natural cubic spline), sex, survey round, schooling, household wellbeing quintile (index rebuilt without the gas stove), rural residence, region, Indigenous language, diagnosed diabetes and smoking. Adjusted %: predictive margin. PR: prevalence ratio against clean fuel; CI: confidence interval. Stove type (item h0110) among households whose main fuel is firewood or charcoal: a gas or electric stove (possible use of several fuels) or another, unspecified stove reported despite solid main fuel; an open fire or oven with a chimney or hood, or a closed oven with a chimney; an open fire or oven without a chimney or hood. Within-solid-fuel ratios are linear contrasts of the same models. Global tests are Wald tests of all groups; the trend test enters clean fuel and the three solid-fuel groups as a score (0–3). p values are not adjusted for multiplicity. n is unweighted. A dash indicates not applicable. Pooled weights divide each annual weight by the number of pooled rounds in which the state had records; the weights of adults aged 60 years or older who answered the functioning section in 2023 and 2024 were rescaled. ENSANUT: Encuesta Nacional de Salud y Nutrición (National Health and Nutrition Survey).

## Subgroups and sensitivity analyses

The PRs for any difficulty seeing were 0.98 (95% CI 0.89 to 1.08) in women and 1.09 (95% CI 0.91 to 1.30) in men (p=0.259 for interaction) and close to 1 in urban and rural areas (Figure 4; Appendices, Table 15). The product term with age group gave p=0.008, with PRs of 0.96 (95% CI 0.85 to 1.08) at 40–59 years and 1.13 (95% CI 1.00 to 1.29; lower limit 0.996) at 60 years or older; when Model 2 was fitted separately in each age group, the PRs were 1.02 (95% CI 0.88 to 1.17) and 1.07 (95% CI 0.93 to 1.22). For a lot of difficulty or inability to see, the ratio of the two age-group PRs pointed in the same direction but was imprecise (1.25, 95% CI 0.83 to 1.89). The other tests of interaction gave p≥0.14, and none was corrected for multiplicity. The PR for glasses use was below 1, with a confidence interval excluding 1, in every subgroup.

**Figure 4:**
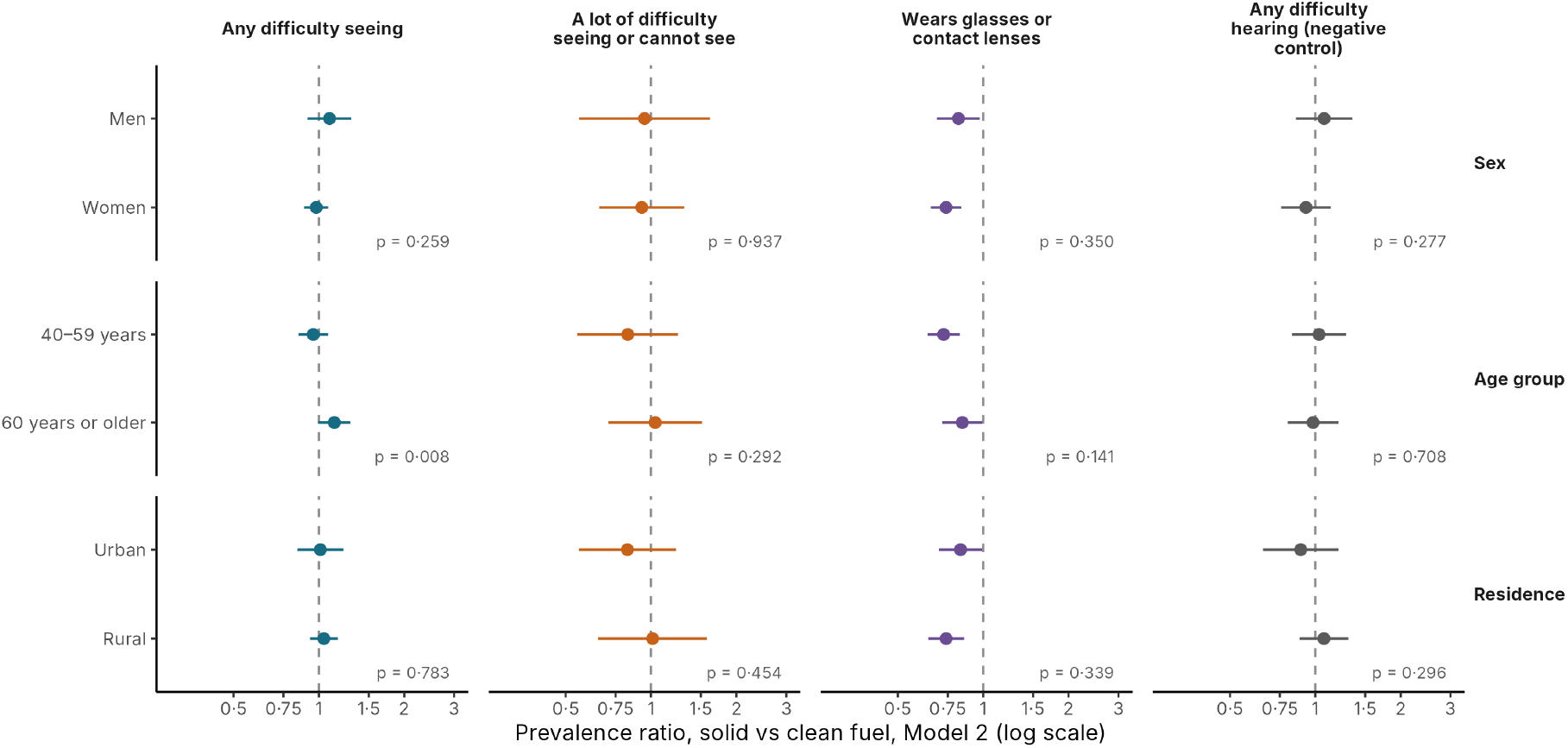
Solid vs clean cooking fuel by sex, age group and residence. Prevalence ratios (logarithmic scale) with 95% confidence intervals from Model 2 with fuel nested within the modifier; p values are Wald tests of interaction, not adjusted for multiplicity. Model 2 covariates: age (natural cubic spline), sex, survey round, schooling, household wellbeing quintile (index rebuilt without the gas stove), rural residence, region, Indigenous language, diagnosed diabetes and smoking.

Across the sensitivity analyses, the PRs for any difficulty seeing ranged from 0.99 to 1.08, all with confidence intervals including 1 and upper limits of up to 1.19; for a lot of difficulty or inability to see, they ranged from 0.93 to 1.24, and for glasses use from 0.71 to 0.79, all with confidence intervals below 1 (Appendices, Table 16 and Figure 5). Using the INSP quintile, which includes the gas stove, gave a PR for any difficulty seeing of 1.03 (95% CI 0.92 to 1.15); omitting the index gave 1.08 (95% CI 0.98 to 1.19); and adding the 2021 round, without the index, which was not released for 2021, gave 1.08 (95% CI 0.99 to 1.17). Tests for heterogeneity between rounds did not provide clear evidence of differences (all p≥0.11), although the round-specific estimates were imprecise (Appendices, Table 17). The E-values for glasses use were 1.90 for the estimate and 1.59 for the confidence limit. In all 18 groups defined by sex and five-year age band, the crude prevalence of difficulty seeing was higher in solid-fuel than in clean-fuel households, and glasses use was lower in 17 of them (Appendices, Figure 6). Coefficients of all terms of Model 2 are given in the Appendices (Table 18).

## Discussion

### Principal findings

About one in 10 Mexican adults aged 40 years or older in the survey lived in a household that cooked mainly with firewood or charcoal. These adults reported more difficulty seeing and hearing than adults in clean-fuel households, but both differences narrowed after accounting for schooling and household wellbeing. After adjustment, there was no clear association with difficulty seeing and no gradient by stove type, place of cooking or sleeping where the household cooks, and the upper confidence limit of the adjusted PR for any difficulty seeing was 1.15. Glasses use was lower in solid-fuel households, and the difference persisted after adjustment.

### Comparison with other studies

Our adjusted estimate is lower than the pooled odds ratio of 1.70 for vision impairment in the 2025 meta-analysis [4]. Outcome definitions differ, but at a baseline prevalence of about 34%, as observed here, an odds ratio of 1.70 corresponds to a PR of about 1.37, above the upper confidence limit of our estimate (1.15). The attenuation with socioeconomic adjustment resembles that in the Longitudinal Ageing Study in India, where the excess of measured vision impairment fell from 9.0 percentage points (adjusted for age and sex) to 3.2 percentage points [7]; our confidence interval for the adjusted difference (−2.8 to 4.9 percentage points) does not exclude a residual excess of that size. Our result is also compatible with the imprecise estimate for cataract in the Mexican sample of a multicountry survey [4]. Larger associations have been reported in Chinese cohorts, with an odds ratio of 1.17 for incident cataract after long-term solid-fuel use [5] and a hazard ratio of 1.38 for incident self-reported distance vision impairment [6]. In those cohorts, solid fuels included coal and the outcomes were incident events, whereas we compared current fuel use with current difficulty.

Our measures could miss a modest association with lens opacity. The Washington Group item captures functional difficulty rather than cataract: in population-based surveys, the “some difficulty or worse” cut-off for seeing identified 67% to 85% of people with clinical distance vision impairment, with a specificity of 77% to 80% [30]. Difficulty seeing after 40 years of age also reflects uncorrected refractive error and presbyopia [21], which dilute any contribution of cataract, and cataract surgery removes the lens opacity that smoke may have caused. The exposure was the household’s current main fuel, not lifetime or personal exposure, and stacking of firewood with LP gas is common in Mexico [10]. Such misclassification could attenuate associations. Even so, there was no gradient with an open fire without a chimney, cooking indoors or sleeping where the household cooks, all of which are expected to raise exposure [11]; in a Chinese cohort, too, associations with cataract were similar with and without ventilated stoves [5].

Associations with cataract have been reported mainly in women in some studies [5,8], whereas studies of reported or measured vision impairment found no larger association in women [6,7], as in our data. The larger PR among adults aged 60 years or older in the interaction model is compatible with an association that increases with cumulative exposure. However, its confidence interval included 1, the contrast was smaller when Model 2 was fitted separately by age group, the pattern for marked difficulty was imprecise, and the comparison was one of 12 interaction tests.

### Difficulty hearing as a negative control

Difficulty seeing and difficulty hearing behaved similarly: both were more common in solid-fuel households when only age, sex and survey round were considered, and both differences were largely attenuated by schooling and household wellbeing. This suggests that the crude excesses mainly reflect the socioeconomic position of solid-fuel users, consistent with the strong social patterning of wood use in Mexico [13]. Cataract accounts for only part of the difficulty seeing reported after 40 years of age, so even an effect on the lens of the size estimated for cataract would raise any difficulty seeing much less, and the confidence interval of the ratio of the two PRs cannot exclude such a diluted effect. A negative control assumes that the two outcomes share their confounders [16], but access to glasses affects reported seeing and not hearing. In addition, in validation surveys the hearing item showed lower and more variable sensitivity for clinical impairment than the seeing item (44% to 83% vs 67% to 85%) [30], which could move a hearing association toward the null. The absence of an association with the negative control does not exclude unmeasured confounding of the primary outcome [16], and solid-fuel use has been associated with hearing loss in Chinese cohorts [23], so a true effect on both senses masked by misclassification cannot be ruled out. The higher prevalence of difficulty hearing in some small groups of solid-fuel households, without a consistent pattern, is more likely to reflect imprecision or residual confounding than an effect of smoke on hearing.

### Glasses use

The most consistent finding was the lower use of glasses: after adjustment, adults in solid-fuel households were 22% less likely to wear them (PR 0.78), and the PR was below 1, with a confidence interval excluding 1, in every subgroup and pooled sensitivity analysis. After adjustment, this shortfall was not accompanied by more reported difficulty seeing, which could reflect lower demands on near vision, different thresholds for reporting difficulty, or lower need; the need for correction was not measured. Before adjustment, adults in solid-fuel households reported more difficulty seeing and wore glasses about half as often. Unaddressed presbyopia and refractive error are the largest components of the vision impairment that could have been prevented or has yet to be addressed [21]. Cooking with firewood or charcoal may also mark deprivation beyond the dwelling characteristics and goods captured by the wellbeing index. The E-values (1.90 for the estimate and 1.59 for the confidence limit) indicate that residual confounding of moderate strength could explain the shortfall. Associations of that size are seen for measured characteristics: in Model 2, glasses use was 1.95 times as common with upper secondary or higher schooling as with no schooling, and schooling differed sharply by fuel.

## Implications

Self-reported difficulty seeing should not be used to attribute vision loss to household air pollution in Mexico without careful control of socioeconomic position. The measured visual acuity, media opacity and red-reflex assessments of the 2025 ENSANUT module will allow cataract to be studied directly [15] and, combined with the household items used here, could test whether lens opacity is more frequent with open fires. Meanwhile, households that cook with solid fuel, which can be identified with a single survey question, could be prioritized for refractive services. The transition to clean cooking is justified regardless of these results by its other health benefits [1,2].

## Strengths and limitations

This study used four pooled rounds of a national survey with harmonized household and adult items, accounted for the complex survey design, reproduced published household estimates, rebuilt the wellbeing index to avoid adjusting for an item close to the exposure, analyzed a negative control outcome jointly with the primary outcome, and included several sensitivity analyses. It also has limitations. First, the design is cross-sectional and the exposure is current: households that recently switched to LP gas carry past exposure into the reference group, which would bias estimates toward the null. Second, vision was reported, not measured, and the item neither identifies cataract nor distinguishes distance from near vision. Third, difficulty seeing refers to vision with glasses for wearers and without them for everyone else; because far fewer adults in solid-fuel households wore glasses, the outcome is closer to uncorrected vision in that group than in clean-fuel households. Fourth, only the main fuel is recorded, not personal exposure, duration of use or who cooks, and the stove category “other” is unspecified. Fifth, the wellbeing index includes dwelling materials that may relate to kitchen ventilation, so adjustment could absorb part of the exposure; however, the PR without the index was also close to 1 (1.08, 95% CI 0.98 to 1.19), and residual confounding by unmeasured deprivation could act in the opposite direction. Sixth, in 2023 and 2024, one half of adults aged 60 years or older were not asked the seeing items by design, and the weights of the other half were rescaled, which assumes that assignment was unrelated to the outcomes. Finally, Sinaloa was absent from two rounds, although results were similar in the 31 states with records in every round.

## Conclusions

Among adults aged 40 years or older in Mexico, the excess of self-reported difficulty seeing in households that cook mainly with firewood or charcoal narrowed after adjustment for socioeconomic position, and so did the excess of the negative control outcome, difficulty hearing. There was no clear gradient by stove type or kitchen arrangements. These findings suggest that crude comparisons mainly reflect the socioeconomic disadvantage of households that use solid fuel, although this outcome measure cannot exclude a modest effect of smoke on the lens.

Glasses use remained lower in solid-fuel households after adjustment, suggesting that these households could be prioritized for refractive services. Studies with measured visual acuity and lens examination are needed to establish whether solid-fuel cooking contributes to cataract in Mexico.

## Data Availability

All data produced in the present study are available upon reasonable request to the authors.

## Acknowledgments

Martín Roberto Basile-Alvarez and Enrique C. Guerra contributed equally to this work and should be considered co-first authors. Data availability: de-identified public-use ENSANUT Continua 2021–2025 data are available from the Instituto Nacional de Salud Pública (https://ensanut.insp.mx/). The article and its appendices report the estimates, and the authors plan to release the analysis code and the definitions of derived variables. Use of artificial intelligence: the authors used cloud-based AI-assisted tools (ChatGPT, OpenAI, San Francisco, CA, USA; primarily GPT-5.6 Sol) to support writing and readability and to generate code for parts of the analysis, and Claude (Anthropic, San Francisco, CA, USA) for language editing. These tools were not used to generate the research question or study design, interpret the findings, or formulate the conclusions. All AI-assisted output was directed, reviewed, tested and revised by the authors, who take full responsibility for the content of the manuscript.

